# Longitudinal analysis of serum neutralization of SARS-CoV-2 Omicron BA.2, BA.4 and BA.5 in patients receiving monoclonal antibodies

**DOI:** 10.1101/2022.08.12.22278699

**Authors:** Timothée Bruel, Karl Stéfic, Yann Nguyen, Donatella Toniutti, Isabelle Staropoli, Françoise Porrot, Florence Guivel-Benhassine, William-Henry Bolland, Delphine Planas, Jérôme Hadjadj, Lynda Handala, Cyril Planchais, Matthieu Prot, Etienne Simon-Lorière, Emmanuel André, Guy Baele, Lize Cuypers, Luc Mouthon, Hugo Mouquet, Julian Buchrieser, Aymeric Sève, Thierry Prazuck, Piet Maes, Benjamin Terrier, Laurent Hocqueloux, Olivier Schwartz

## Abstract

The emergence of novel Omicron lineages, such as BA.5, may impact the therapeutic efficacy of anti-SARS-CoV-2 neutralizing monoclonal antibodies (mAbs). Here, we evaluated the neutralization and ADCC activity of 6 therapeutic mAbs against Delta, BA.2, BA.4 and BA.5 isolates. The Omicron sub-variants escaped most of the antibodies but remained sensitive to Bebtelovimab and Cilgavimab. Consistent with their shared spike sequence, BA.4 and BA.5 displayed identical neutralization profiles. Sotrovimab was the most efficient at eliciting ADCC. We also analyzed 121 sera from 40 immunocompromised individuals up to 6 months after infusion of 1200 mg of Ronapreve (Imdevimab + Casirivimab), and 300 or 600 mg of Evusheld (Cilgavimab + Tixagevimab). Sera from Ronapreve-treated individuals did not neutralize Omicron subvariants. Evusheld-treated individuals neutralized BA.2 and BA.5, but titers were reduced by 41- and 130-fold, respectively, compared to Delta. A longitudinal evaluation of sera from Evusheld-treated patients revealed a slow decay of mAb levels and neutralization. The decline was more rapid against BA.5. Our data shed light on the antiviral activities of therapeutic mAbs and the duration of effectiveness of Evusheld pre-exposure prophylaxis.

## Intro

Nine months after its emergence, the SARS-CoV-2 Omicron lineage outcompeted previous variants of concerns (VOCs). Sublineages with improved transmissibility have replaced the initial Omicron BA.1 strain. As of July 2022, Omicron was composed of 5 main sublineages, BA.1, BA.2, BA.3, BA.4 and BA.5^1,2^, harboring numerous mutations compared to the ancestral wuhan strain ^1,2^. The BA.1 strain has 34 mutations in its spike, associated with antibody escape^3–14^, CD8 T cell evasion^15^ and modified tropism^16–18^. BA.2 harbors 30 mutations, 21 of which are not present in BA.1^2^. The BA.4 and BA.5 sublineages share the same spike sequence, which differs from BA.2 by three mutations (including one reversion) in the Receptor Binding Domain (RBD) and one deletion in the N-terminal domain (NTD)^1^. The BA.5 sublineage was dominant in many countries in July 2022^19^.

Several monoclonal antibodies (mAbs) targeting the spike protein of SARS-CoV-2 are used in therapeutic, pre-exposure prophylaxis (PrEP) and post-exposure prophylaxis (PEP) settings^20^. Therapeutic administration of mAbs is highly effective, reaching 85% efficacy in preventing Coronavirus disease 19 (COVID-19)-related hospitalization or death^21–23^. Antibody-based prophylaxis also achieves high levels of protection. Ronapreve (Imdevimab + Casirivimab) and Evusheld (Cilgavimab + Tixagevimab) cocktails provide 81% and 77% protection against symptomatic infection, respectively ^24,25^. These successes are mitigated by viral evolution. Omicron variants display considerable escape from mAbs^3,4,6–9,12,14,26–28^. The use of Ronapreve (Imdevimab + Casirivimab) and Sotrovimab was discouraged after BA.1 and BA.2 emergence^29^. It is recommended to inject a double-dose of Evusheld, as serum neutralization is decreased against BA.1 and BA.2 in individuals receiving these antibodies as PrEP^28–31^. Bebtelovimab is similarly effective against ancestral strains and Omicron BA.1 and BA.2^32^, but its access is so far restricted to the United States^33^. Thus, a continuous evaluation of mAbs efficacy against new variants is needed to optimize their utilization.

As other Omicron sublineages, BA.4 and BA.5 escape most neutralizing mAbs^34–41^. The neutralization profile of BA.4 and BA.5 is similar to that of BA.2, with only Cilgavimab and Bebtelovimab being efficient against these strains with high potency^34–38^. Animal models revealed that some mAbs do not only rely on neutralization for therapeutic efficacy^42,43^. Antibodies can trigger effector mechanisms through their fragment crystallizable (Fc) region. These so-called Fc-effector functions mediate killing of infected cells through activation of antibody-dependent cellular cytotoxicity (ADCC) by NK cells, or clearance of viral particles, through macrophages-mediated antibody-dependent phagocytosis (ADP)^44^. Interaction between the Fc region and cognate Fc Receptors (FcR) may also promote inflammation and antibody-dependent enhancement (ADE) of infection^45^. Hence, some therapeutic mAbs were intentionally mutated in the Fc region to abrogate FcR recognition and eliminate putative risk of ADE^20^. This is the case for Cilgavimab and Tixagevimab, which bear a triple mutation (TM) motif (L234F, L235E and P331S). Fc engineering may also modulate neonatal Fc receptor (FcRn) affinity and extend antibody half-life^46^. Such modifications were introduced in Sotrovimab (M428L and N434S, called LS), Cilgavimab and Tixagevimab (M252Y, S254Y and T256E, called YTE). Therefore, the therapeutic activity of antibodies is the sum of neutralization potency, Fc-effector functions and bio-disponibility.

Here, we evaluated the neutralization and ADCC activity of 6 therapeutic mAbs against BA.4 and BA.5 isolates. We also analyzed serum neutralization from 40 immunocompromised individuals up to 6-months post-infusion of Ronapreve (Imdevimab + casirivimab) and Evusheld (Cilgavimab + Tixagevimab).

## Results

### In vitro neutralization of BA.4 and BA.4

We first investigated the sensitivity of two authentic isolates of BA.4 and BA.5 to neutralization by mAbs. We selected 6 antibodies that are either used in patients (Cilgavimab, Tixagevimab and Bebtelovimab) or that were withdrew because of Omicron escape (Sotrovimab, Casirivimab and Imdevimab). We used the commercial formulation except for Bebtlovimab, which is not available in France. We also tested Ronapreve (Imdevimab + Casirivimab) and Evusheld (Cilgavimab + Tixagevimab) cocktails. As controls, we included Delta and BA.2 strains^26,28^. We used our S-Fuse neutralization assay^26,28,47,48^, based on syncytia formation, to quantify infection via a green fluorescent protein (GFP) split system^26,28,46,47^.

The IC50 of 4 out of the 6 mAbs (Sotrovimab, Tixagevimab, Casirivimab and Imdevimab) were higher for BA.4 and BA.5 than Delta **(figure 1a and table 1)**. Tixagevimab and Casirivimab lacked neutralization in the range of concentrations tested **(figure 1a and table 1)**. Sotrovimab and Imdevimab remained active but lost potency. As compared to Delta, Sotrovimab was 15- and 17-fold less potent against BA.4 and BA.5, respectively. The increase in IC50s was higher for Imdevimab, 110-fold and 86-fold against BA.4 and BA.5, respectively. Imdevimab remained more potent than Sotrovimab against both strains (IC50 of 265 and 996 ng/ml, for BA.4, and 208 and 1088 ng/ml for BA.5, respectively) **(figure 1a and table 1)**. Importantly, Cilgavimab and Bebtelovimab displayed no or only minimal changes as compared to delta, and remained highly potent against BA.4 and BA.5. When compared to BA.2, BA.4 and BA.5 display slightly improved neutralization by Imdevimab (4.2- and 5.3-fold) and Sotrovimab (9- and 8.3-fold) **(figure 1a and table 1)**. We also analyzed the combination of Cilgavimab and Tixagevimab (Evusheld by Astrazeneca) and Casirivimab and Imdevimab (Ronapreve by Regeneron). Both displayed a drop in potency compared to Delta, which was less marked for Evusheld (BA.4: 10.4-fold and BA.5: 9-fold) than Ronapreve (BA.4: 330-fold and BA.5: 350-fold) **(figure 1a and table 1)**.

**Figure 1:**
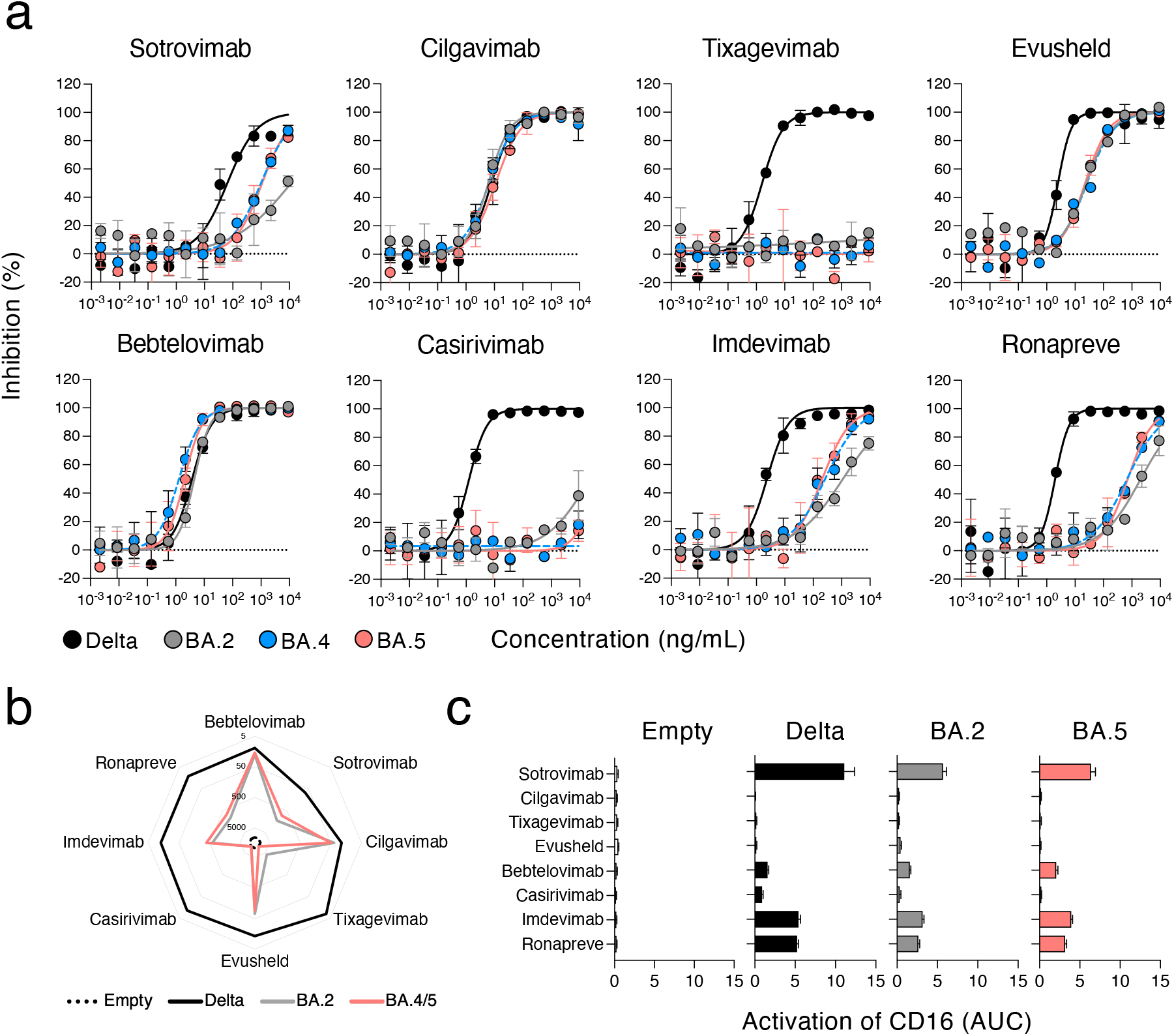
Neutralization and antibody-dependent cellular cytotoxicity of Omicron BA.4 and BA.5 by therapeutic mAbs. **a**. Neutralization curves of mAbs. Dose–response analysis of the neutralization by the indicated antibodies and by Evusheld, a combination of cilgavimab and tixagevimab, and Ronapreve, a combination of casirivimab and imdevimab. Data are mean ± s.d. of 2 independent experiments. The IC50 values for each antibody are presented in Table 1. **b**. mAbs binding at the surface of Raji cells stalby expressing the indicated spikes. Depicted are EC50, calculated with are curve fitting the % of mAbs positive cells measured by flow cytometry against antibody C° in limiting-dilutions. The EC50 values for each antibody are also presented in Table 1. **c**. Activation of the CD16 pathway as a surrogate of the capacity of each mAbs to elicit antibody-dependent cellular cytotoxicity (ADCC). The area under curve of a dose-response analysis of CD16 activation by each mAbs against each SARS-CoV-2 variant is depicted.

**Table 1 :** Summary of in vitro antiviral activities of therapeutic mAbs against Delta, BA.2 and BA.4.

|  | Neutralization |  |  |  | Binding |  |  | ADCC |  |  |
| --- | --- | --- | --- | --- | --- | --- | --- | --- | --- | --- |
|  | Delta | BA.2 | BA.4 | BA.4 | Delta | BA.2 | BA.4 | Delta | BA.2 | BA.4/5 |
| Sotrovimab | 64.18 | >9000 | 996 | 1088 | 72.3 | 1429 | 848.2 | ++++ | +++ | +++ |
| Cilgavimab | 7.9 | 6.1 | 6.5 | 11 | 22 | 39 | 49.3 | - | - | - |
| Tixagevimab | 1.6 | >9000 | >9000 | >9000 | 7.8 | 4226 | >9000 | - | - | - |
| Evusheld | 2.5 | 24.7 | 26.1 | 22.6 | 13.5 | 72.9 | 92.3 | - | - | - |
| Bebtelovimab | 3.8 | 4.5 | 1.3 | 2 | 12.2 | 20.2 | 17.4 | + | + | + |
| Casirivimab | 1.3 | >9000 | >9000 | >9000 | 11.1 | >9000 | >9000 | + | - | - |
| Imdevimab | 2.4 | 1120 | 265 | 208 | 12.8 | 610 | 391 | ++ | + | + |
| Ronapreve | 2 | 1985 | 660 | 700 | 12.8 | 1106 | 739 | ++ | ++ | ++ |
|  | IC50 (ng/mL) |  |  |  | EC50 (ng/mL) |  |  |  |  |  |

Overall, our data reveal a large escape of therapeutic mAbs by Omicron BA.4 and BA.5. BA.2. BA.4 and BA.5 have a similar profile of neutralization by these mAbs. Cilagavimab and Bebtelovimab remain fully active against these variants.

### Antibody binding to BA.4/5 spike and induction of ADCC

Next, we evaluated the capacity of these mAbs to bind to BA.4 and BA.5 spike (referred to as BA.4/5 spike) and trigger ADCC. We assessed antibody binding by flow cytometry, using Raji cells stably expressing the BA4/5 spike. As controls we included Delta, BA.2 spike and cells transduced with an empty vector. To confirm spike expression and compare the various cells lines, we stained the cells with Bebtlovimab, which neutralized Delta, BA.2, BA.4 and BA.5 with a similar potency. All cell lines showed >90% of spike expression with similar median fluorescence intensity (MFI) across variants **(supplementary figure 1b)**. No binding was observed with either Raji-Empty cells or the isotype control (mGO53) **(figure 1b)**.

We analyzed the binding of the therapeutic mAbs and their combinations against these cells. We performed limiting dilution tests to calculate exact effective concentrations 50% (EC50) **(figure 1b, table 1 and supplementary figure 1b)**. All antibodies bound the Delta spike, with EC50 <100 ng/ml. This was expected given their neutralizing potency against this strain. Binding profiles were generally similar between BA.2 and BA.4/5, with both spikes displaying high level of escape compared to Delta **(figure 1b)**. Bebtelovimab and Cilgavimab displayed similar binding levels. Tixagevimab and Casirivimab did not recognize the BA.4/5 spike, even at a high concentration (10*μ*g/mL). Sotrovimab and Imdevimab recognized the BA.4/5 spike, with a loss of potency compared to delta (11.7- and 30.5-fold, respectively) **(figure 1b, table 1 and supplementary figure 1b)**.

Then, we investigated the capacity of these mAbs to trigger ADCC. We used a surrogate assay that measures the activation of the CD16 pathway. We previously demonstrated that this assay correlates to killing of infected cells by primary NK cells^48^. We tested the antibodies by limiting dilution. We measured the area under the curve (AUC) to depict the ADCC capacity of the mAbs against each viral spike **(figure 1c, table 1 and supplementary Figure 1c)**. As expected, none of the mAbs elicited CD16 activation against the Raji-Empty cells. CD16 activation was detectable against Raji-Delta, -BA.2- and -BA.4/5 cells. Sotrovimab was the most efficient mAb, regardless of the viral strain, albeit the AUC was reduced against BA.2 and BA.4/5 compared to Delta **(Figure 1c, table 1 and supplementary Figure 1c)**. Cilgavimab and Tixagevimab alone or in the Evusheld cocktail did not activate ADCC, in line with the mutations in their Fc domain that decrease binding to FcR **(figure 1c, table 1 and supplementary Figure 1c)**. Bebtelovimab induced similar levels of ADCC activation against all strains, yet at low levels **(figure 1c table 1 and supplementary Figure 1c)**. Imdevimab and Casirivimab, alone or in the Ronapreve cocktail displayed intermediate levels of activation **(figure 1c table 1 and supplementary Figure 1c))**.

Overall, our results indicate that BA.4/5 avoid antibody recognition and ADCC activation by most of therapeutic mAbs tested. Sotrovimab is the most efficient ADCC inducer, and Cilgavimab and Tixagevimab lack ADCC activity.

### Serum neutralization of BA.5 in individuals receiving mAbs

Next, we investigated antibody levels and neutralization in sera from 40 immunocompromised individuals who received by intra-muscular injection either 300mg (n=29) or 600mg (n=11) of Evusheld as PrEP. Patients were sampled prior to and at a median of 26 (range 10-40) or 37 (range 14-48) days after the single or double dose, respectively. Among the 29 individuals who received 300mg of Evusheld, 17 previously received Ronapreve as PrEP. The last injection of Ronapreve occurred at a median of 35 days (range 29-49) before the first Evusheld injection. Two out of the 11 individuals who received 600 mg of Evusheld also received Ronapreve. However, the injections were spaced by >160 days, which is ∼5-times above the half-life of Ronapreve^49^. Thus, we considered them as naive at the time of Evusheld administration. Participants were included in the study in two places, the Centre Hospitalier Regional (CHR) in Orléans (France; n=8) or the Hôpital Cochin in Paris (France; n=32). Most of the patients were female (n=28), were diagnosed with ANCA-Associated vasculitis (n=26) and treated with rituximab as immunosuppressive therapy (n=31). A complete description of the patients’ characteristics is provided in **table 2**. The 8 individuals recruited at the CHR Orléans were longitudinally sampled every month as part of an ongoing prospective cohort.

**Table 2 :** Characteristics of the patients included in the study.

|  | "Orléans" cohort | "Cochin" Group | Total (%) |
| --- | --- | --- | --- |
| patient characteristics |  |  |  |
| n | 8 | 32 | 40 (100) |
| Gender ♀ | 6 | 22 | 28 (70) |
| Gender ♂ | 2 | 10 | 12 (30) |
| obesity | 3 | 1 | 4 (10) |
| Diseases |  |  |  |
| Rheumatoid arthritis | 5 | 2 | 7 (17.5) |
| Kidney graft | 2 | 0 | 2 (5) |
| myelodysplasia | 1 | 0 | 1 (2.5) |
| ANCA-Associated Vasculitis | 0 | 26 | 26 (65) |
| polychondritis | 0 | 1 | 1 (2.5) |
| lupus | 0 | 1 | 1 (2.5) |
| systemic sclerosis | 0 | 1 | 1 (2.5) |
| cryoglobulinemic vasculitis | 0 | 1 | 1 (2.5) |
| medications |  |  |  |
| Rituximab (anti-CD20) | 5 | 26 | 31 (77.5) |
| Infliximab (anti-TNF) | 0 | 1 | 1 (2.5) |
| Mepolizumab (anti-IL-5) | 0 | 1 | 1 (2.5) |
| Prednisone | 4 | 10 | 14 (35) |
| Mycophenolate Mofetil | 2 | 1 | 3 (7.5) |
| Methotrexate | 0 | 3 | 3 (7.5) |
| 5-azacytidine | 1 | 0 | 1 (2.5) |
| Tacrolimus | 1 | 0 | 1 (2.5) |
| Cyclosporin | 1 | 0 | 1 (2.5) |
| Cyclophosphamide | 0 | 1 | 1 (2.5) |
| Vaccines doses |  |  |  |
| 2 | 0 | 1 | 1 (2.5) |
| 3 | 5 | 25 | 30 (75) |
| 4 | 3 | 6 | 9 (22.5) |
| previous COVID-19 | 0 | 8 | 8 (20) |
| PrEP |  |  |  |
| Ronapreve | 3 | 17 | 20 (50) |
| Evusheld 300mg | 8 | 21 | 29 (72.5) |
| Evusheld 600mg | 0 | 11 | 11 (27.5) |

We analyzed the 40 individuals before and after infusion of Evusheld. We categorized the patients into 5 groups: naïve (before treatment), Ronapreve, Ronapreve+Evusheld (who received Ronapreve followed by 300mg of Evulsheld), Evusheld (who received 300mg) and Evusheldx2 (who received 600mg). We first measured the levels of anti-spike IgGs in binding antibody units (BAU/mL) using the S-Flow assay. As compared to the naive group, sera containing mAbs display a sharp increase in anti-S IgG (median of 38 vs 3449, 3591, 1323 and 2623 BAU/mL, for Ronapreve, Ronapreve+Evusheld, Evusheld and Evusheldx2, respectively) **(figure 2b)**. We then investigated serum neutralization against Delta, BA.2 and BA.5 with the S-Fuse assay. We tested sera in limiting dilutions to calculate titers as effective dilution 50% (ED50). We did not include BA.4, as its profile of neutralization is identical to BA.5. Untreated individuals (naïve group) did not neutralize the three strains, except for one patient who slightly neutralized Delta. Infusion of mAbs dramatically increased Delta neutralization,with increases from 552 to 2484-fold compared to the naive group. Individuals who received Ronapreve neutralized BA.2 and BA.5 at low levels (1.5- and 1.2-fold increase over naive individuals; non-significant). The groups that included Evusheld neutralized BA.2 and BA.5 at levels significantly higher than the naive and Ronapreve groups **(figure 2b)**. Of note, neutralization titers were higher in the Evusheldx2 group with all variants tested (16585 vs 23772, 992 vs 1908 and 511 vs 539, against Delta, BA.2 and BA.5, respectively), without reaching statistical significance. Neutralization titers tended to be lower against BA.5 than BA.2, but this difference was significant only for individuals who received 300 mg of Evusheld (1549 vs 489; p= 0.0228) **(supplementary figure 2)**.

**Figure 2:**
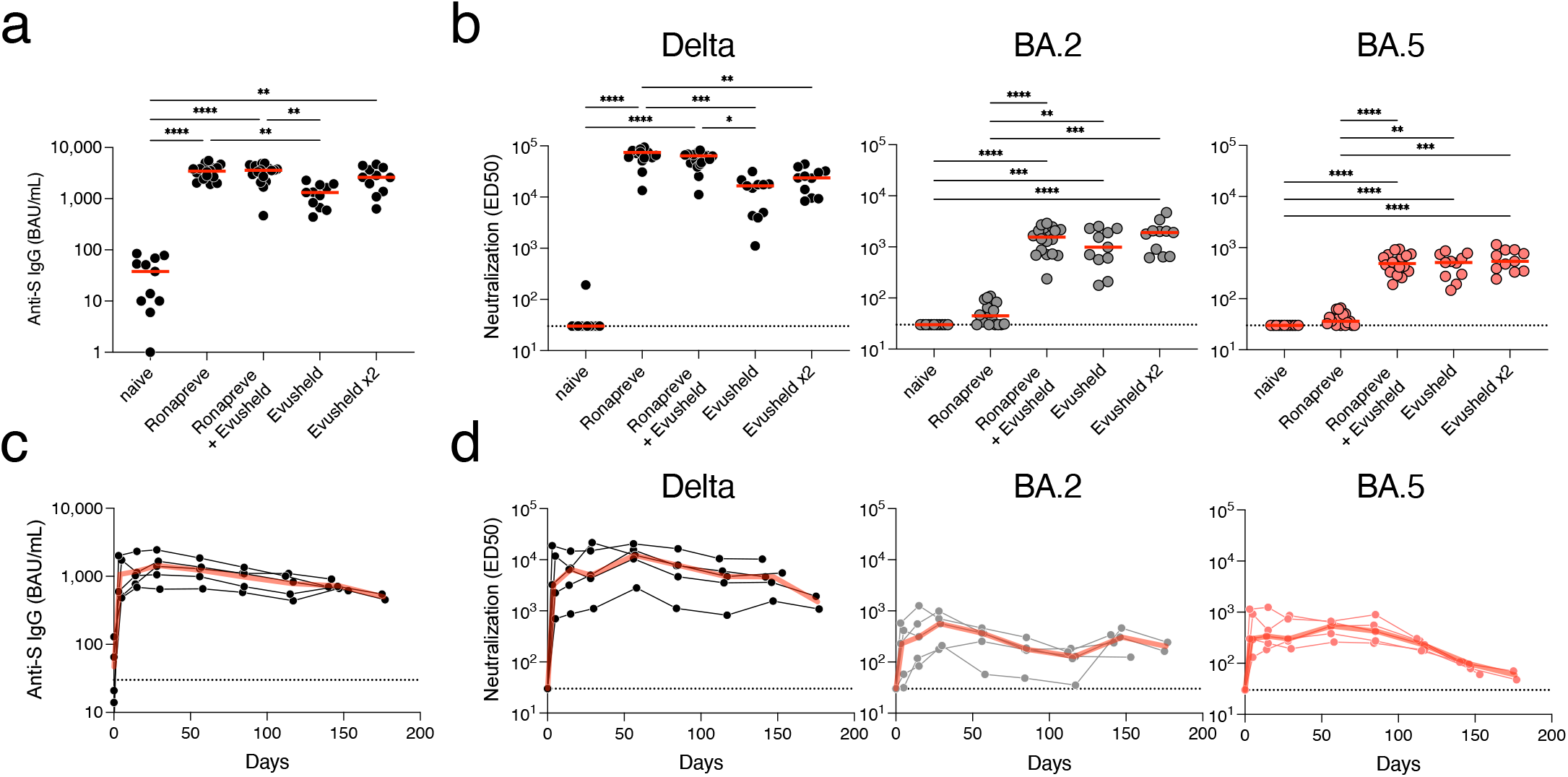
Antibody levels and neutralization of delta, BA.2 and BA.5 in sera of immunocompromised individuals receiving mAbs. **a**. Anti-S IgGs were measured using the flow cytometry-based S-Flow assay in sera of individuals before PrEP (naive; n = 11), treated with Ronapreve (n = 18), treated with 300mg (n = 11) or 600mg (n=11) of Evusheld, or treated with both Ronapreve and 300mg of Evusheld (n = 18). Indicated are the binding antibody unit (BAU) per mL (BAU/mL) of anti-S IgGs. Two-sided Kruskall–Wallis test with Dunn’s multiple comparison correction. Each dot is an individual. Red bars indicate median. **b**. Serum neutralization of Delta and Omicron BA.2 and BA.5 in the same individuals as in (a). Indicated are Effective dilution 50% (ED50; titres) as calculated with the S-Fuse assay. Two-sided Kruskall–Wallis test with Dunn’s multiple comparison correction. Each dot is an individual. Red bars indicate median. **c**. Longitudinal measurement of anti-S levels in 5 immunocompromised individuals who initiated an Evusheld PrEP with no history of Ronapreve. All individuals and sampling points are depicted (black lines and dots). The red lines indicate the median. Indicated are the binding antibody unit (BAU) per mL (BAU/mL) of anti-S IgGs. **d**. Sero-neutralization of Delta and Omicron BA.2 and BA.5 in the same individuals as in (c). Indicated are Effective dilution 50% (ED50 ; titres) as calculated with the S-Fuse assay. Two-sided Kruskall–Wallis test with Dunn’s multiple comparison correction. Each dot is an individual. Red bars indicate median.

Overall, these data show that the serum neutralization activity of individuals receiving Ronapreve and Evusheld as PrEP is decreased against BA.2 and BA.5. The diminution is less marked with Evusheld-than Ronapreve-treated individuals.

### Kinetics of serum neutralization up to 6-months after infusion of Evusheld

Longitudinal sampling was performed for 8 out of the 40 individuals (for patients’ characteristics see **table 2**). The serum samples were available up to 186 days post administration of Evusheld (300mg). We investigated anti-S IgG levels and neutralization **(Figure 2c,d and supplementary Figure 3)**. We first analyzed 5 patients who were naive at the time of Evusheld injection (the 3 others were previously under Ronapreve PrEP) **(Figure 2c,d)**. Antibody levels peaked at 28 (range 28-30) days after Evusheld administration, with a median of 1400 (range 646-4014) BAU/mL. Anti-S IgG then slowly decreased, reaching 500 (range 452-548) BAU/mL at 176 (range 175-177) days post-administration **(Figure 2c)**. Neutralization of Delta, BA.2 and BA.5 mirrored anti-S levels, showing a sharp increase upon administration and a steady decrease until month 6 **(Figure 2d)**. Neutralization of Delta remains consistently higher than that of BA.2 and BA.5, in line with our other observations **(figure 1a, 2b and supplementary figure 2)**. After almost 6 months of follow up, the five patients who received Evusheld harbored detectable levels of neutralization against the tested strains. The neutralization levels against the two Omicron subvariants were low at 6 months (ED50 of 1503, 202 and 59, for Delta, BA.2 and BA.5, respectively) **(Figure 2d)**. We also analyzed the 3 individuals who received Ronapreve prior to Evusheld **(supplementary figure 3)**. Together, their profile was similar to those who only received Evusheld. However, they consistently harbored higher levels of anti-S and neutralization titers in the first two months, suggesting a disappearance of Ronapreve, but a maintenance of Evusheld, as expected given the longer half-life of Evusheld than Ronapreve.

Overall, these results show that a single administration of Evusheld allowed serum neutralization of BA.5 for 6 months, with reduced titers compared to Delta.

## Discussion

We show here that BA.4 and BA.5 escape neutralization by most therapeutic mAbs, in line with previous reports^34–41^. Some antibodies remain effective. Bebtelovimab is the most potent, followed by Cilgavimab. Tixagevimab and Casirivimab lost any neutralizing activity and Imdevimab was poorly active against BA.4/BA.5. We observed a slightly higher neutralization of BA.5 by Sotrovimab compared to BA.2. Similar findings were reported^37,38,41^, but others have found decreased or identical neutralization of BA.5 compared to BA.2^34–36,39,40^. This may be explained by the binding domain of Sotrovimab, which recognizes the RBD outside of the receptor binding motif (RBM)^50^. This may render Sotrovimab more susceptible to experimental variations, such as the use of different target cells, viral isolates or pseudotypes.

In addition to our in vitro evaluation of mAbs neutralization, we analyzed the sera of immunocompromised individuals receiving Ronapreve or Evusheld as PrEP. In line with our in vitro observation, Ronapreve-treated individuals barely neutralized Omicron sublineages. Evusheld-treated individuals had detectable neutralization against BA.2 and BA.5, albeit decreased compared to Delta. We observed a trend for a higher neutralization of BA.2 than BA.5 in individuals receiving Evusheld. Longitudinal evaluation in 8 patients showed a slightly faster decay of antibody responses against BA.5. This difference between BA.2 and BA.5 may be explained by the loss of BA.5 binding and neutralization by Tixagevimab. This decrease may be negligible when the Evusheld antibodies are tested alone, but more visible in the serum. How serum components might affect Evusheld potency against BA.5 will deserve further investigations. Cilgavimab may also be slightly less potent against BA.4/5 than BA.2, as reported by others^35,36,38^. The difference of BA.2 and BA.5 serum neutralization in Evusheld-treated individuals and the observation of a faster antibody decay stress the need for a booster dose of mAbs after 6 months, as currently recommended. It may be of great interest to evaluate the impact of an earlier booster dose of Evusheld to compensate for Omicron escape.

There was large inter-individual variability in neutralization and antibody levels after mAb administration. Recent work demonstrated an impact of the body mass index (BMI) on antibody levels after Evusheld injection, with high BMI associated to low titers^30^. This is consistent with the unique recommended dosage of Evusheld (initially 300mg and then 600 mg). In our study, we observed a non-significant trend for higher titers in individuals receiving 600 mg and no association to BMI. This lack of significance is likely due to the small number of individuals tested and to additional factors accounting for the inter-individual’s variability. An investigation of a larger cohort previously demonstrated a significant increase in antibody levels in individuals received 600 mg as compared to 300 mg^31^. It will be interesting to evaluate whether adapting the dose to BMI may homogenize the response to Evusheld PrEP and improve its efficacy.

We also tested the binding and ADCC capacity of these mAbs. Binding correlated to neutralization, but not to ADCC. The most potent antibody to activate ADCC against Omicron sublineages was Sotrovimab, even if its neutralization IC50 was relatively high compared to other antibodies. This ADCC activity may help understand why Sotrovimab remains clinically active against BA.2, despite its very limited neutralization^51^. Similarly, it has been reported that non-neutralizing antibodies capable of mediating Fc-effector functions display some efficacy in animal models^52^. It may be worth examining whether a combination of Sotrovimab and Evusheld or Bebtelovimab may improve therapeutic efficacy of the mAbs.

Our study has limitations. First, our sample size is small, precluding the analysis of patient characteristics associated to high serum neutralization titers. Whether biological sex, age, ongoing medication, or underlying condition modulate bio-disponibility of mAbs remains open questions. Second, we did not have access to mucosal samples. Systemic levels of antibodies are known to be key to protect the lung and prevent severe COVID-19, whereas mucosal mAb levels may correlate with protection from infection. Our study was also limited to BA.4 and BA.5, and we did not analyze the sensitivity of other Omicron subvariants, such as BA.2.75 ^53–55^. We did not have access to the medical formulation of Bebtelovimab^33^. Fc-effector functions are influenced by the method of antibody preparation, the isotype, Fc glycosylation and mutations. Our observation that Bebtelovimab is a poor ADCC inducer deserves to be confirmed using the medical formulation.

In conclusion, we provide here an in-depth evaluation of the efficacy of therapeutic mAb and serum from mAb-treated patients against Omicron sublineages. The predominant BA.5 variant remains sensitive to Evusheld, but the decay of the serum neutralizing activity in treated individuals is accelerated, compared to previously circulating variants.

## Methods

### Cohorts

Individuals under Evusheld PreP were recruited in the French cities of Orléans and Paris (CHR d’Orléans and Hôpital Cochin). The Neutralizing Power of Anti-SARS-CoV-2 Serum Antibodies (PNAS) cohort is an ongoing prospective, monocentric, longitudinal, observational cohort clinical study aiming to describe the kinetics of neutralizing antibodies after SARS-CoV-2 infection or vaccination (ClinicalTrials.gov identifier: NCT05315583). The cohort takes place in Orélans, France and enrolled immunocompromised individuals receiving Evusheld PreP. This study was approved by the Est II (Besançon) ethical committee. At enrollment, written informed consent was collected, and participants completed a questionnaire that covered sociodemographic characteristics, clinical information and data related to anti-SARS-CoV-2 vaccination. Blood sampling was performed on the day of Evusheld infusion and after 3 days, 15 days and then every months. The ‘Cochin’ cohort is a prospective, monocentric, longitudinal, observational clinical study (NCT04870411) enrolling immunocompromised individuals with rheumatic diseases, aiming at describing immunological responses to COVID-19 vaccine in patients with autoimmune and inflammatory diseases treated with immunosuppressants and/or biologics. Ethics approval was obtained from the Comite de Protection des Personnes Nord-Ouest II. Leftover sera from usual care were used from these individuals in the setting of the local biological samples collection (RAPIDEM). A written informed consent was collected for all participants. None of the study participants received compensation.

### Viral strains

All strain were isolated from a nasopharyngeal swab using Vero E6 cells (ADCC: CRL-1586™) tested negative for mycoplasma. The Delta and Omicron BA.2 strains were previously described^26,28^. BA.4 and BA.5 strains were isolated from Belgian and French patients, respectively. BA.4 was isolated and sequenced by the NRC UZ/KU Leuven (Belgium). BA.5 was isolated from a 67-year-old female patient. On May 15, she experienced mild and unspecific symptoms, she tested positive for COVID-19 using a lateral flow assay. Due to pre-existing conditions (polymyalgia rheumatica), she presented at the hospital on 17/05, where a nasal swab was collected for PCR testing and sequencing, identifying BA.5. Her COVID-19 symptoms remain mild (arthralgia and cough). Both patients provided informed consent for the use of the biological materials. The sequences of the isolates were deposited on GISAID immediately after their generation, with the following Delta ID: EPI_ISL_2029113; Omicron BA.2 GISAID ID: EPI_ISL_10654979. Omicron BA.5: EPI_ISL_13660702. viral stocks were titrated in limiting dilution on Vero E6 cells and on S-Fuse cells.

### mAbs

Bamlanivimab, Casirivimab, Etesevimab, Imdevimab, Cilgavimab, Tixagevimab and Sotrovimab were provided by CHR Orleans. Bebtelovimab was produced as previously described^28^.

### Cell lines

Raji cells (ATCC CCL-86) were grown in complete RPMI medium (10% Fetal Calf Serum (FCS), 1% Penicillin/Streptomycin (PS)). 293T cells (ATCC CRL-3216) and U2OS cells (ATCCa HTB-96) were grown in complete DMEM medium (10% FCS, 1% PS). U2OS stably expressing ACE2 and the GFPsplit system (GFP1-10 and GFP11; S-Fuse cells) were previously described (ref). Blasticidin (10 mg/mL) and puromycin (1 mg/mL) were used to select for ACE2 and GFPsplit transgenes expression, respectively. Raji cells stably expressing the SARS-CoV-2 Spike protein of Delta, BA.2 and BA.4/5 (GenBank: QHD43416.1, UJP23605.1 and UPN16705.1) were generated by lentiviral transduction and selection with puromycin (1 mg/mL). Absence of mycoplasma contamination was confirmed in all cell lines with the Mycoalert Mycoplasma Detection Kit (Lonza). All cell lines were cultured at 37°C and 5% CO2.

### Anti-spike antibody binding and serology

Circulating levels of anti-S antibodies were measured with the S-Flow assay. This assay uses 293T cells stably expressing the spike protein (293T spike cells) and 293T control cells as control to detect anti-spike antibodies by flow cytometry^56^. In brief, the cells were incubated at 4 °C for 30 minutes with sera (1:300 dilution) in PBS containing 0.5% BSA and 2 mM EDTA. Cells were then washed with PBS and stained with an anti-human IgG Fc Alexa Fluor 647 antibody (109-605-170, Jackson Immuno Research). After 30 minutes at 4 °C, cells were washed with PBS and fixed for 10 minutes using 4% PFA. A standard curve with serial dilutions of a human anti-spike monoclonal antibody (mAb48) was acquired in each assay to standardize the results as a binding Unit (BU). Data were acquired on an Attune NxT instrument using Attune NxT software version 3.2.2 (Life Technologies) and analyzed with FlowJo version 10.7.1 software (see Extended Data Fig. 4 for gating strategy). The sensitivity is 99.2% with a 95% confidence interval of 97.69–99.78%, and the specificity is 100% (98.5– 100%)40. To determine BAU ml−1, we analyzed a series of vaccinated (n = 144), convalescent (n = 59) samples and World Health Organization international reference sera (20/136 and 20/130) on S-Flow and on two commercially available ELISAs (Abbott 147 and Beckmann 56). Using this dataset, we performed a Passing–Pablok regression, which shows that the relationship between BU and BAU ml−1 is linear, allowing calculation of BAU ml−1 using S-Flow data^57^. The binding mAbs to Delta, BA.2 and BA.4/5 spikes was assessed using Raji cells stably expressing these spikes. Stainings were performed at the indicated concentration of mAbs and following the S-Flow protocol, except that antibodies were biotinylated and revealed with a streptavidin conjugated to AlexaFluor647 (Life Technologies ; dilution 1:400).

### S-Fuse neutralization assay

U2OS-ACE2 GFP1-10 or GFP11 cells, also termed S-Fuse cells, become GFP+ when they are productively infected by SARS-CoV-2. Cells tested negative for mycoplasma. Cells were mixed (ratio 1:1) and plated at 8 × 103 per well in a μClear 96-well plate (Greiner Bio-One). The indicated SARS-CoV-2 strains were incubated with serially diluted mAb or sera for 15 minutes at room temperature and added to S-Fuse cells. The sera were heat-inactivated for 30 minutes at 56 °C before use. Eighteen hours later, cells were fixed with 2% paraformaldehyde (PFA), washed and stained with Hoechst (dilution 1:1,000, Invitrogen). Images were acquired with an Opera Phenix high-content confocal microscope (PerkinElmer). The GFP area and the number of nuclei were quantified using Harmony software version 4.9 (PerkinElmer). The percentage of neutralization was calculated using the number of syncytia as value with the following formula: 100 × (1 − (value with serum − value in ‘non-infected’) / (value in ‘no serum’ − value in ‘non-infected’)). Neutralizing activity of each serum was expressed as the ED50. ED50 values (in μg ml−1 for mAbs and in dilution values for sera) were calculated with a reconstructed curve using the percentage of the neutralization at the different concentrations. We previously reported correlations between neutralization titers obtained with the S-Fuse assay and both pseudovirus neutralization and microneutralization assays^58,59^.

### Antibody-dependent cellular cytotoxicity reporter assay

ADCC was quantified using the ADCC Reporter Bioassay (Promega) as previously described^48^. Briefly, 5×10^4^ Raji stably expressing the indicated spikes were co-cultured with 5×10^4^ Jurkat-CD16-NFAT-rLuc cells in presence or absence of pre-pandemic or COVID-19 sera at the indicated dilution. Luciferase was measured after 18 hours of incubation using an EnSpire plate reader (PerkinElmer). ADCC was measured as the fold induction of Luciferase activity compared to the ‘‘no serum’’ condition.

### Statistical analysis

No statistical methods were used to predetermine sample size. The experiments were not randomized, and the investigators were not blinded. Flow cytometry data were analyzed with FlowJo version 10 software. Calculations were performed using Excel 365 (Microsoft). Figures were drawn on Prism 9 (GraphPad Software). Statistical analysis was conducted using GraphPad Prism 9. Statistical significance between different groups was calculated using Kruskall–Wallis test with Dunn’s multiple comparisons, Friedman tests with Dunn’s multiple comparison correction and Spearman non-parametric correlation test. All tests were two-sided.

### Data availability

All data supporting the findings of this study are available within the article or from the corresponding authors upon reasonable request without any restrictions.

## Acknowledgments

We thank the European Health Emergency Preparedness and Response Authority (HERA) for supporting the work being done at Institut Pasteur and UK Leuven. We thank the patients who participated to this study. We thank members of the Virus and Immunity Unit for discussions and help. We thank N. Aulner and the UtechS Photonic BioImaging (UPBI) core facility (Institut Pasteur), a member of the France BioImaging network, for image acquisition and analysis. The Opera system was co-funded by Institut Pasteur and the Région Ȋle-de-France (DIM1Health). We thank F. Peira, V. Legros and L. Courtellemont for their help with the cohorts. UZ Leuven, as national reference center for respiratory pathogens, is supported by Sciensano, which is gratefully acknowledged. We thank Hélène Péré and David Veyer for their help in sequencing viral strains and helpful discussions.

## Author contribution

Experimental strategy and design: T.B and O.S.

Laboratory experiments: T.B, D.T, I.S, F.P, F.G-B, W-H.B, D.P, M.P, E.S-L and J.B

Cohort management and clinical research: Y.N, J.H, L.M, A.S, T.P, B.T and L.H.

Viral strains and antibodies: K.S, L.H, C.P, E.A, G.B, L.C, H.M,

Manuscript writing: T.B. and O.S.

Manuscript editing: T.B, Y.N, W-H.B, B.T, L.H and O.S.

## Supplementary Figure Legends

**Supplementary figure 1:**
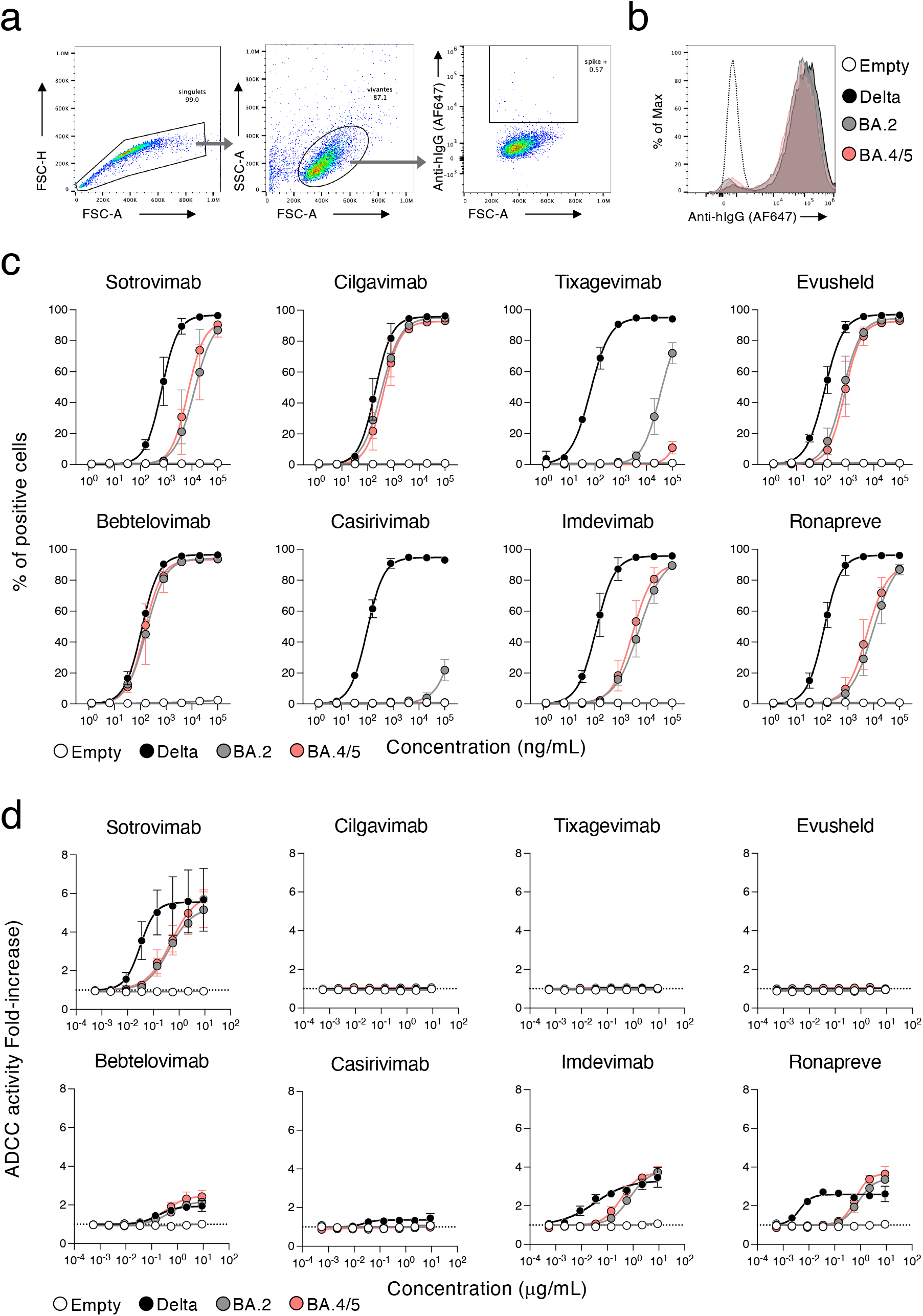
Capacity of therapeutic antibodies to bind and elicit ADCC against the BA.4/5 spike. **a**. Gating strategy of the binding assay. Raji cells stably expressing and empty transgene were incubated with bebtelovimab conjugated to biotin (200ng/mL), stained with a streptavidin coupled to AlexaFluor 647 (AF647) and analyzed by flow-cytometry. A representative example of the gating strategy is shown. **b**. An example of the fluorescence signal obtained with bebtelovimab (200ng/mL) on the Raji cells expressing Delta, Omicron BA.2 and Omicron BA.5 spikes. The Raji empty cells are used as control. **c**. Dose–response analysis of the binding by the indicated antibodies and by Evusheld, a combination of cilgavimab and tixagevimab, and Ronapreve, a combination of casirivimab and imdevimab. The % of mAbs positive cells measured by flow cytometry against antibody C° in limiting-dilutions are depicted. Data are mean ± s.d. of 2 independent experiments. The EC50 values for each antibody are presented in Table 1. **d**. Dose–response analysis of the ADCC activity by the indicated antibodies and by Evusheld, a combination of cilgavimab and tixagevimab, and Ronapreve, a combination of casirivimab and imdevimab. The fold-increase in CD16 activation as compared to the “no Raji” condition is indicated for each concentration of mAb. Data are mean ± s.d. of 2 independent experiments. Area under the curve are scored and summarized in Table 1.

**Supplementary figure 2:**
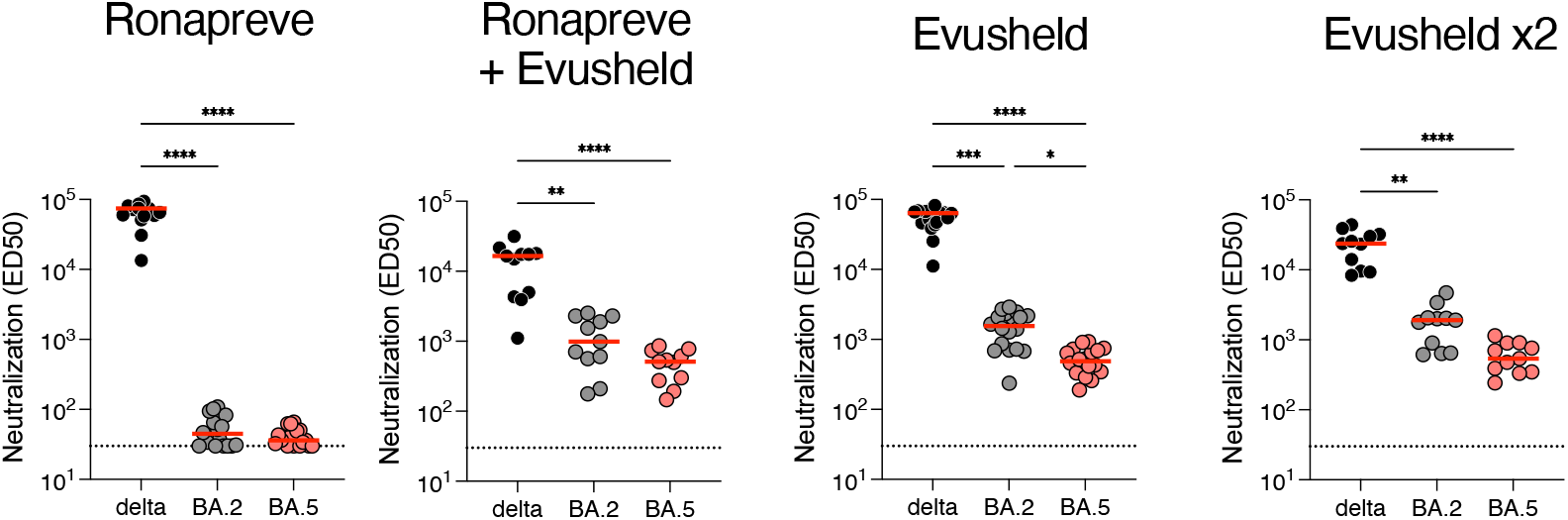
Antibody levels and neutralization of delta, BA.2 and BA.5 in sera of immunocompromised individuals receiving mAbs. Serum neutralization of Delta and Omicron BA.2 and BA.5 in the same individuals as in Figure 2. Indicated are Effective dilution 50% (ED50; titres) as calculated with the S-Fuse assay. Two-sided Kruskall–Wallis test with Dunn’s multiple comparison correction. Each dot is an individual. Red bars indicate median.

**Supplementary figure 3:**
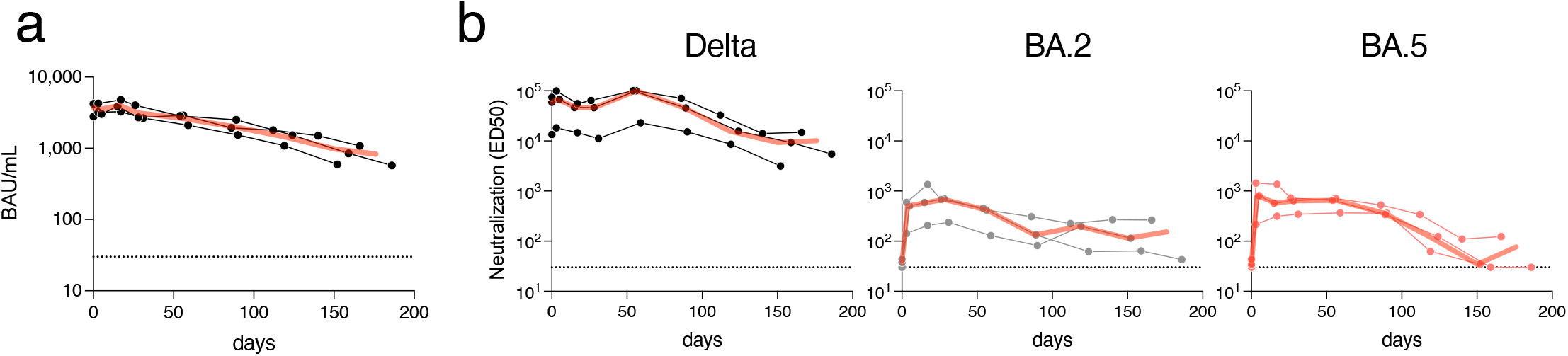
Longitudinal evaluation of antibody levels and neutralization in 3 individuals who switched from Ronapreve to Evusheld. Serum neutralization of Delta and Omicron BA.2 and BA.5 in the 3 individuals who switched from Ronapreve to Evusheld for their SARS-CoV-2 PrEP. Indicated are Effective dilution 50% (ED50 ; titres) as calculated with the S-Fuse assay. Two-sided Kruskall–Wallis test with Dunn’s multiple comparison correction. Each dot is an individual. Red bars indicate median.

